# Stroke Mortality During the War in Ukraine

**DOI:** 10.64898/2026.10.05.26364812

**Authors:** Oleksandr Topachevskyi, Ian J. Stewart, Evgeniy Gonchar, Dmytro Lebedynets, Jeffrey T. Howard

**Author notes:** Corresponding Author: Jeffrey T. Howard, University of Texas at San Antonio, One UTSA Circle, San Antonio, Texas 78249,.

## Abstract

**Background:** Mounting evidence shows war directly and indirectly harms civilian health. As the largest conflict since World War II, the war in Ukraine severely impacts civilian survival. Stroke—a leading cause of death in Ukraine—represents a key population-level vulnerability to full-scale war. We assessed the war’s impact on population-level stroke mortality.

**Methods:** This retrospective cohort study evaluated patients diagnosed with hemorrhagic or ischemic stroke admitted to Ukrainian civilian hospitals from April 1, 2020, through December 31, 2024. Using data from the Ukrainian nationwide electronic health record system, we compared pre-war and post-war stroke mortality. We used multivariate proportional hazards regression with propensity matching to test if in-hospital mortality increased. Excess deaths were estimated by multiplying the pre-post mortality rate difference by the post-period population in person-days.

**Results:** Among 577,501 total individuals, we found that the crude in-hospital stroke mortality rate increased by 117 (95% CI: 94.2-139.9) per 100,000 person-days in the post-war period. In propensity-matched analyses, the mortality risk was 5% (HR=1.05; 95% CI: 1.03-1.06; p<0.001) higher in the post-war period. Mortality risk varied by age, sex, admission type and stroke type. Patients with hemorrhagic stroke had 179% (HR=2.79; 95% CI: 2.75-2.83; p<0.001) higher risk of death compared to ischemic stroke patients.

**Conclusion:** In the first study to assess stroke mortality at the population level in Ukraine after the start of the full-scale war, we found that in-hospital mortality increased in the post-war period with an estimated 3,944 excess stroke deaths.

## INTRODUCTION

There is growing evidence that war has both direct and indirect health effects on military and civilian populations^1^. The majority of the evidence is from studies of US and coalition military service members serving in the global war on terror in Iraq and Afghanistan have shown that military exposures during war, such as environmental contaminant exposure, combat exposure and battle and non-battle injuries, are associated with a wide range of long-term health problems, including post-traumatic stress disorder (PTSD)^2–4^, depression^3,4^, anxiety^3,4^, substance abuse, cardiovascular disease (CVD)^5–10^, cancer^11,12^, diabetes^6,7^, chronic kidney disease^6^, respiratory diseases^8^ and premature mortality^13^. Studies of US veterans of the Gulf War report similar findings^14^.

Most research on the impact of war on civilian populations has focused on morbidity and mortality from war-related injuries^15^. Fewer studies have investigated the indirect impact of war on civilian health outcomes. A systematic review of war impacts on civilian health found consistent evidence linking exposure to war and increased CVD and CVD mortality, while the results for stroke and stroke mortality were mixed^16^. Several studies found evidence supporting an association between war exposure and hypertension^17,18^, CVD^19^, CVD mortality^17,20^, stroke^17,20,21^ and stroke mortality^17,20^. In a more recent study of civilian health impacts following the September 11, 2001 terrorist attacks showed exposed civilians had elevated risk of cardiovascular death similar in magnitude to first-responders^22^, which was consistent with findings in a study of Iraqi civilians and war-related cardiovascular mortality during the Iraq conflict^23^.

Stroke mortality has been the 3^rd^ leading cause of death in Ukraine as recently as 2021^24^, just prior to Russia’s launch of its full-scale invasion of Ukraine, which triggered the largest war in Europe since World War II. The scale of the ongoing conflict suggests that civilians may be suffering from significant health burdens as an indirect result of the war. A recent study of stroke incidence and mortality in Kyiv, Ukraine found that both incidence and mortality increased after the start of the full-scale war^25^. However, another study of the Ukrainian city of Ivano-Frankivsk found that there was no difference in stroke outcomes in the post-war period^26^. To-date there has been no assessment of the impact of the war on stroke mortality at the national level. Our goal was to test the hypothesis that in-hospital mortality from stroke increased after the start of the full-scale Russian invasion of Ukraine and to estimate the number of excess stroke deaths during the post-war period.

## METHODS

### Study Design and Participants

A retrospectively determined cohort study of in-hospital mortality for patients admitted to hospitals in Ukraine with cerebrovascular (stroke) events from April 1, 2020 through December 31, 2024. Inclusion criteria included hospital admission during the study period a primary diagnosis of International Classification of Disease, Australian Modification, version 10 (ICD-10-AM) of I61 nontraumatic intracerebral hemorrhage (hemorrhagic stroke) or I63 cerebral infarction (ischemic stroke). The research was determined to be exempt from review by the University of Texas at San Antonio institutional review board and conducted in accordance with the Declaration of Helsinki and with applicable Ukrainian and U.S. regulations. This study followed the Strengthening the Reporting of Observational Studies in Epidemiology (STROBE) reporting guideline for cohort studies.

### Data

The data were provided by Ministry of Health of Ukraine. The dataset is a full extract from the nationwide electronic health record (EHR) system named “eHealth”, which was launched in April 2020. The eHealth system contains basic demographics, diagnosis codes, hospital admission type, length of stay, and discharge status.

### Measures

The primary exposure of interest in this study was the onset of the full-scale Russo-Ukrainian War, which was defined as pre-war (before February 24, 2022) or post-war (February 24, 2022, or after). The outcome for the study was measured as in-hospital death (yes/no) and follow-up time for each individual patient was defined as the time to discharge or death, in person days. For aggregate analysis, mortality rates per 100,000 person-days were calculated as the sum number of patients admitted in each period divided by the sum of person-days divided by 100,000. Age was measured as continuous age in years, and sex was measured as male or female. Hospital admission type was categorized as emergency, physician referral, self, transfer from other hospital or missing. The primary diagnosis for each patient was based on 4-digit ICD-10-AM coding for I61.0-I61.9 (hemorrhagic stroke) and I63.0-I63.9 (ischemic stroke).

### Statistical Analysis

For individual level analyses, descriptive statistics are reported as mean and standard deviation (SD) or median interquartile range (IQR) for continuous variables, and count and percent for categorical variables. The MatchIt package was used to perform propensity matching to balance covariates between the two periods. Proportional hazards regression (survival package) was used to assess the association between pre-war/post-war period and time-to-death in both the total sample and the matched sample, with results reported as hazard ratios (HR), 95% confidence intervals (CI) and p values. The proportional hazards assumption was tested using Schoenfeld residuals. Survival curves (ggsurvfit package) were used to visualize time-to-death for the total sample and the matched sample, with 95% CI and the p value for the log-rank test, with follow-up time restricted to 90 days. Excess deaths were estimated by taking the difference between the pre-war and post-war mortality rates per 100,000 person-days for the total sample and multiplying by the total person-days divided by 100,000 and reported with 95% CI. Data were analyzed using R version 4.5.3 (R Core Team).

## RESULTS

A total of 577,501 individual patients were included in the analysis, with a median follow-up time of 9 days (IQR: 6-11) accounting for 5,445,823 person-days (14,920 person-years) of observation, and 96,537 deaths from 2020 to 2024 (**Table 1**). There were 206,499 patients (35.8%) in the pre-war period (prior to 2/24/2022) and 371,002 patients (64.2%) in the post-war period (2/24/2022 and after). Mean age for the sample was 68.1 years (SD=11.8) and 51.3% were male. The two most frequent diagnoses were I63.3-Cerebral infarction due to thrombosis of cerebral arteries (42.5%) and I63.5-Cerebral infarction due to unspecified occlusion or stenosis of cerebral arteries (26.4%) accounted for 69% of all patient diagnoses, which differed between the pre-war (I63.3=37% and I63.5=28%) and post-war (I63.3=45% and I63.5=25%) periods (p<0.001). All variables differed significantly between the pre-war and post-war periods.

**Table 1.**
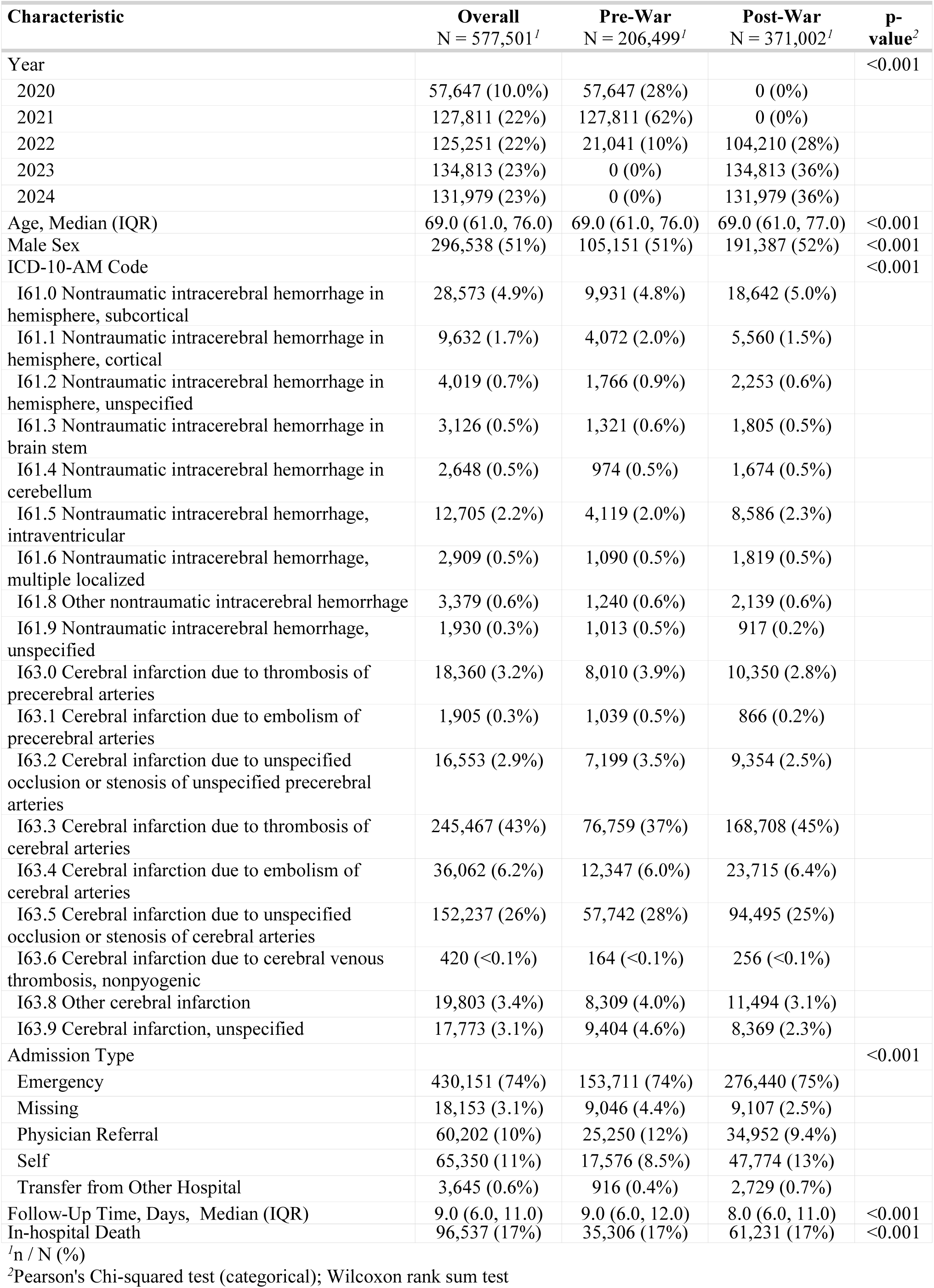
Study Characteristics.

Results of propensity-matching suggest that covariates were well balanced after matching (**Figure 1**). Survival curves show separation between pre-war and post-war periods from 1 to 8 months of follow-up in both the total sample (**Figure 2, panel A**) and the propensity-matched sample (**Figure 2, panel B**) (p<0.001). Although survival curves converge after 8 months, the population at-risk in the pre-war period was 38 in both the total and propensity-matched samples and in the post-war period was 10 in the total sample and 8 in the propensity-matched sample.

**Figure 1.**
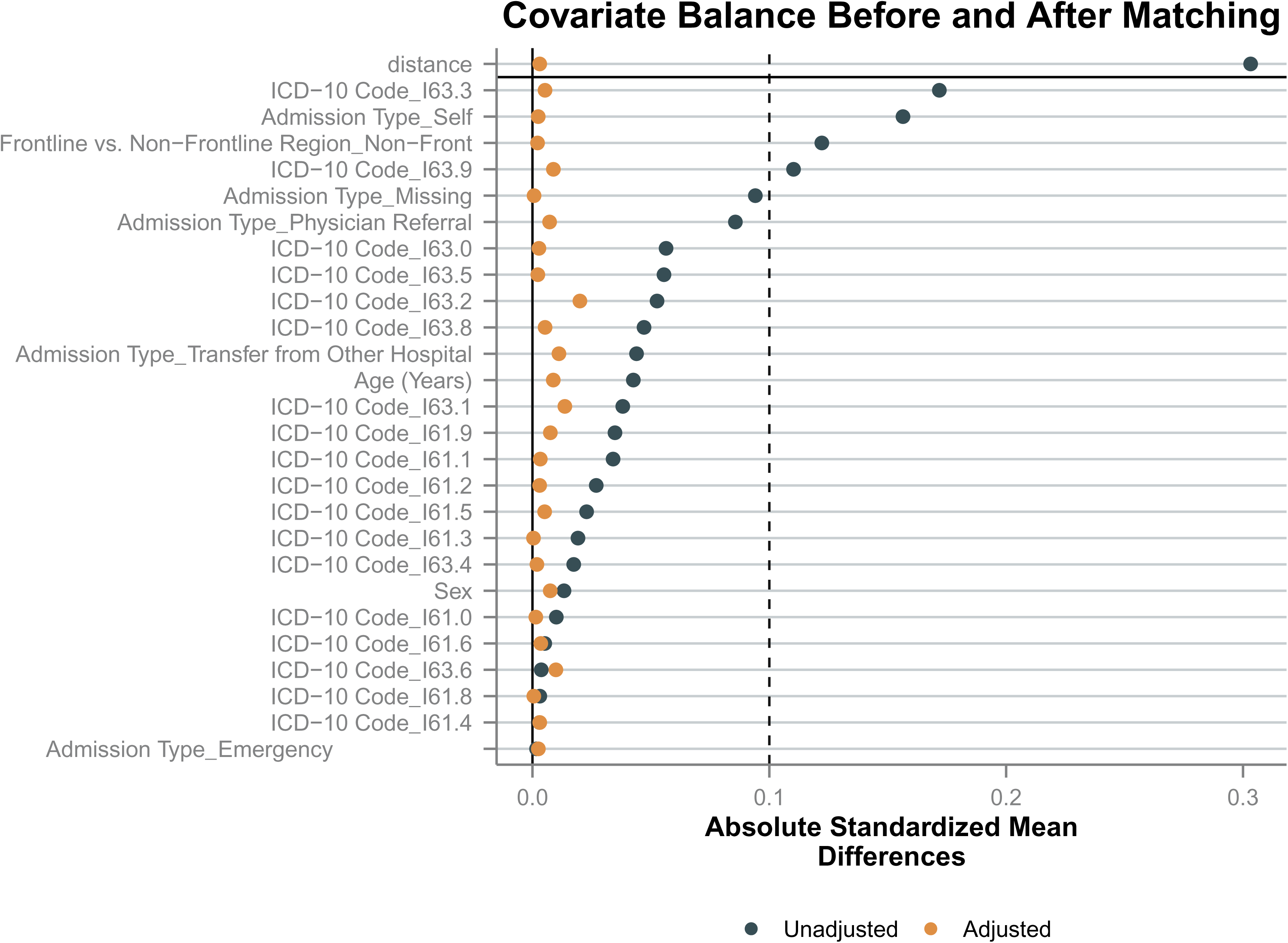
Covariate balance plot before (unadjusted) and after (adjusted) propensity matching.

**Figure 2.**
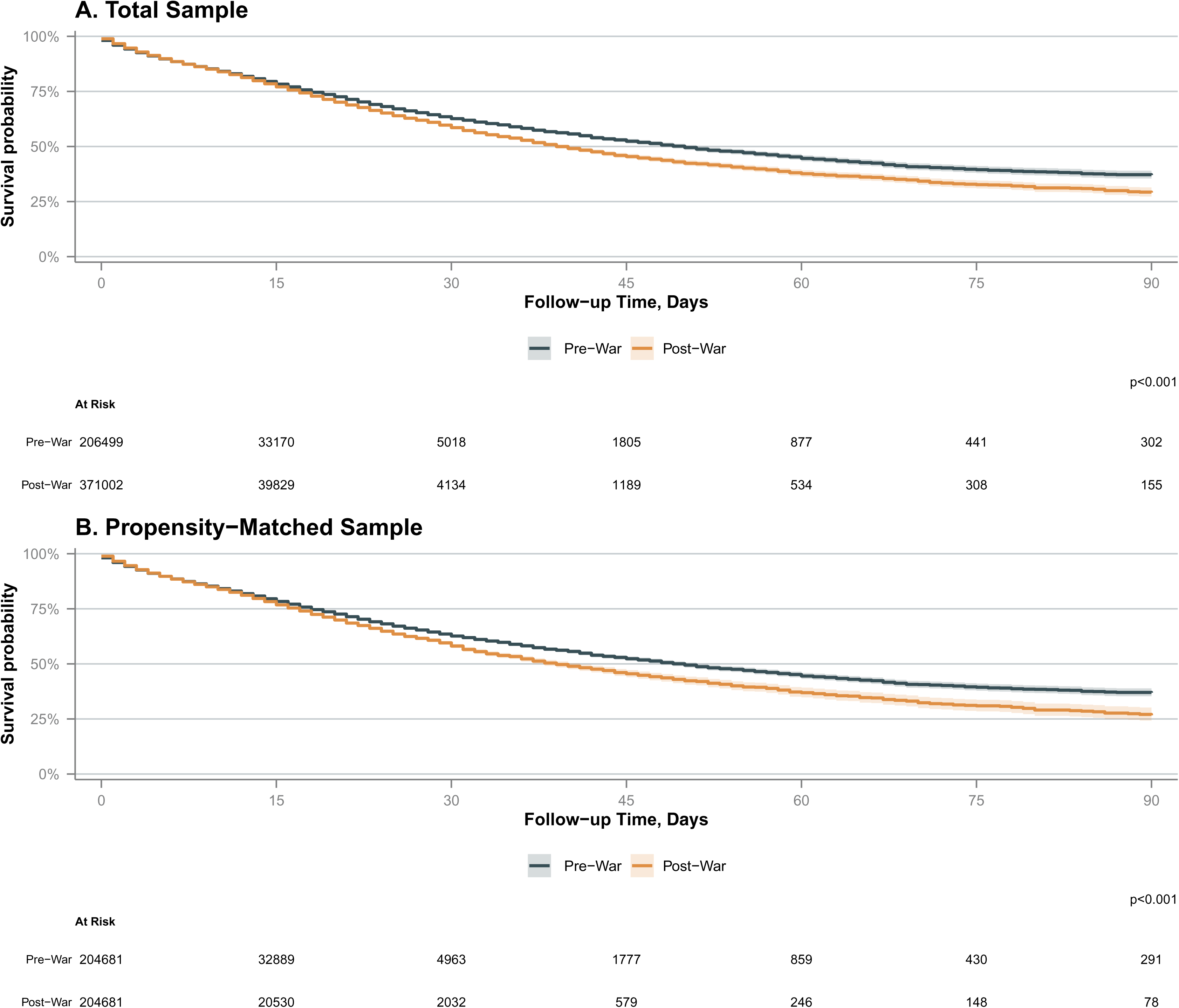
Survival curves comparing stroke in-hospital mortality before and after the onset of the Russo-Ukrainian War in the total sample (A) and propensity-matched sample (B), restricted to 90-day follow-up.

Results of multivariable proportional hazards regression on the total sample indicate a 2% (HR=1.02; 95% CI: 1.01-1.04; p=0.001) increase in the risk of death in the post-war period compared with the pre-war period (**Table 2**). In the propensity-matched sample, the risk of death was 5% higher in the post-war period (HR=1.05; 95% CI: 1.03-1.06; p<0.001). In addition, older age (HR=1.03; 95% CI: 1.03-1.03; p<0.001), male sex (HR=1.05; 95% CI: 1.04-1.07; p<0.001) and admission type of emergency (HR=2.45; 95% CI: 2.37-2.53; p<0.001), self-admit (HR=1.24; 95% CI: 1.19-1.29; p<0.001) and transfer from other hospital (HR=3.35; 95% CI: 3.12-3.60; p<0.001), compared with physician referral, were also associated with increased risk of death. Risk of death also varied significantly by stroke type. Patients with hemorrhagic stroke (I61) had 179% (HR=2.79; 95% CI: 2.75-2.83; p<0.001) higher risk of death compared to ischemic stroke patients, with specific diagnoses of I61.6 nontraumatic intracerebral hemorrhage multiple localized (HR=3.05; 95% CI: 2.90-3.21; p<0.001), I61.3 nontraumatic intracerebral hemorrhage brain stem (HR=2.89; 95% CI: 2.74-3.05; p<0.001), I61.5 nontraumatic intracerebral hemorrhage intraventricular (HR=2.61; 95% CI: 2.53-2.69; p<0.001) representing the three diagnoses with the highest mortality risk. Review of Schoenfeld residuals did not indicate violation of the proportional hazards assumption.

**Table 2.** Results of proportional hazards regression of stroke mortality.

| Characteristic | Total Sample<br>(N=577,501) |  |  | Propensity-Matched<br>Sample<br>(N=409,362) |  |  |
| --- | --- | --- | --- | --- | --- | --- |
|  | HR | 95% CI | p-value | HR | 95% CI | p-value |
| Period |  |  |  |  |  |  |
| Pre-War | — | — |  | — | — |  |
| Post-War | 1.02 | 1.01, 1.04 | 0.001 | 1.05 | 1.03, 1.06 | <0.001 |
| Age | 1.03 | 1.03, 1.03 | <0.001 |  |  |  |
| Sex, male vs. female | 1.05 | 1.04, 1.07 | <0.001 |  |  |  |
| Admission Type |  |  |  |  |  |  |
| Physician Referral | — | — |  |  |  |  |
| Emergency | 2.45 | 2.37, 2.53 | <0.001 |  |  |  |
| Self | 1.24 | 1.19, 1.29 | <0.001 |  |  |  |
| Transfer from Other Hospital | 3.35 | 3.12, 3.60 | <0.001 |  |  |  |
| Missing | 1.55 | 1.47, 1.64 | <0.001 |  |  |  |
| Stroke Type |  |  |  |  |  |  |
| I63 - Ischemic | — | — |  |  |  |  |
| I61 - Hemorrhagic | 2.79 | 2.75, 2.83 | <0.001 |  |  |  |
| Specific ICD-10-AM Codes |  |  |  |  |  |  |
| I61.0 Nontraumatic intracerebral hemorrhage in hemisphere, subcortical | — | — |  |  |  |  |
| I61.1 Nontraumatic intracerebral hemorrhage in hemisphere, cortical | 0.97 | 0.93, 1.01 | 0.17 |  |  |  |
| I61.2 Nontraumatic intracerebral hemorrhage in hemisphere, unspecified | 1.64 | 1.55, 1.73 | <0.001 |  |  |  |
| I61.3 Nontraumatic intracerebral hemorrhage in brain stem | 2.89 | 2.74, 3.05 | <0.001 |  |  |  |
| I61.4 Nontraumatic intracerebral hemorrhage in cerebellum | 0.91 | 0.84, 0.99 | 0.02 |  |  |  |
| I61.5 Nontraumatic intracerebral hemorrhage, intraventricular | 2.61 | 2.53, 2.69 | <0.001 |  |  |  |
| I61.6 Nontraumatic intracerebral hemorrhage, multiple localized | 3.05 | 2.90, 3.21 | <0.001 |  |  |  |
| I61.8 Other nontraumatic intracerebral hemorrhage | 1.43 | 1.34, 1.52 | <0.001 |  |  |  |
| I61.9 Nontraumatic intracerebral hemorrhage, unspecified | 2.39 | 2.22, 2.56 | <0.001 |  |  |  |
| I63.0 Cerebral infarction due to thrombosis of precerebral arteries | 0.62 | 0.59, 0.65 | <0.001 |  |  |  |
| I63.1 Cerebral infarction due to embolism of precerebral arteries | 0.72 | 0.65, 0.80 | <0.001 |  |  |  |
| I63.2 Cerebral infarction due to unspecified occlusion or stenosis of unspecified precerebral arteries | 0.47 | 0.44, 0.49 | <0.001 |  |  |  |
| I63.3 Cerebral infarction due to thrombosis of cerebral arteries | 0.52 | 0.50, 0.53 | <0.001 |  |  |  |
| I63.4 Cerebral infarction due to embolism of cerebral arteries | 0.57 | 0.55, 0.59 | <0.001 |  |  |  |
| I63.5 Cerebral infarction due to unspecified occlusion or stenosis of cerebral arteries | 0.50 | 0.49, 0.51 | <0.001 |  |  |  |
| I63.6 Cerebral infarction due to cerebral venous thrombosis, nonpyogenic | 1.06 | 0.86, 1.30 | 0.586 |  |  |  |
| I63.8 Other cerebral infarction | 0.65 | 0.63, 0.68 | <0.001 |  |  |  |
|  | HR | 95% CI | p-value | HR | 95% CI | p-value |
| I63.9 Cerebral infarction, unspecified | 0.78 | 0.75, 0.82 | <0.001 |  |  |  |

Abbreviations: CI = Confidence Interval, HR = Hazard Ratio

The total mortality rate per 100,000 person-days was 1700.3 in the pre-war period and 1817.3 in the post-war period, a difference of 117.0 (95% CI: 94.2-139.9) (**Table 3**). The estimated number of excess deaths in the post-war period was 3,944 (95% CI: 3,174-4,714).

**Table 3.** Excess mortality estimates.

|  | Total Sample<br>N = 577,501 |
| --- | --- |
| Pre-war |  |
| Number of deaths | 35,306 |
| Follow-up time (person-days) | 2,076,516 |
| Mortality rate per 100,000 person-days | 1700.3 |
| Post-war |  |
| Number of deaths | 61,231 |
| Follow-up time (person-days) | 3,369,307 |
| Mortality rate per 100,000 person-days | 1817.3 |
| Excess mortality estimates |  |
| Mortality rate difference (95% CI) | 117.0 (94.2-139.9) |
| Excess deaths (95% CI) | 3,944 (3,174-4,714) |

## DISCUSSION

To our knowledge, this is the first population level study of civilian patients hospitalized for stroke events in Ukraine and the first to assess excess stroke mortality following the onset of the full-scale Russo-Ukrainian War in February 2022. In this retrospective cohort study, we found that in-hospital mortality increased in the post-war period. In propensity-matched analysis, in which covariates were well balanced, the mortality risk was 5% higher in the post-war period than in the pre-war period. In addition, there was an estimated 3,944 excess deaths in the 34 months following the war onset. This equates to 116 excess deaths per month during the post-war period from February 24, 2022 through December 31, 2024. These findings support the hypothesis that war has health impacts on civilian populations beyond war-related traumatic injuries.

Previous studies of stroke incidence and mortality in Ukraine focused on specific cities, such as Kyiv^25^ and Ivano-Frankivsk^26^. Consistent with our findings, both stroke incidence and in-hospital mortality were shown to have increased in the post-war period in Kyiv, compared to the pre-war period^25^. However, Shkoruta et. al.^26^ showed that while stroke incidence increased in the post-war period, stroke mortality did not change meaningfully during the post-war period in Ivano-Frankivsk. One distinct advantage of our study is the fact that it contains data for the total population of Ukraine, rather than a single city or region.

Other recent studies on the impact of the Israel-Gaza war^27–29^ suggest that while overall stroke and cardiovascular incidence may have increased with the onset of the war, mortality did not. Each of these studies had small samples less than 1000 and were all conducted in Israel. No similar population-based studies of the impact of war on stroke or cardiovascular disease in Gaza have been reported. However, one survey-based study in Gaza showed that the most frequently reported chronic health conditions were hypertension, diabetes mellitus and cardiovascular disease and that common issues facing patients with these conditions included forced displacement, food and water insecurity, disease-related complications, lack of access to medication, and lower self-rated quality of life^30^.

While there is mounting evidence from studies of US and UK military veterans of the Global War on Terror that war impacts long-term cardiovascular^5–7,9,10^ and mental health^2,3,5,31^, research on the impact of war on the health of civilians in Afghanistan and Iraq has focused mainly on direct impacts from traumatic injuries sustained during combat operations, rather than impacts on chronic health conditions. However, one study did assess mortality before and after the start of the Iraq war^23^. Consistent with our results, Hagopian et al found that overall mortality and cardiovascular mortality both increased after the onset of the war^23^. A systematic review that included studies from the wars in Croatia, Bosnia, Columbia, Lebanon, Georgia, Sudan, Iraq and Uganda concluded that incidence of cardiovascular and stroke diseases increased during these wars compared to the pre-war period, as did cardiovascular risk factors, such as obesity, alcohol and tobacco use, and hypertension^16^. Associations between war onset and outcomes were most consistent for cardiovascular mortality, broadly, while results for stroke mortality, specifically, were mixed.

While our study cannot determine the specific factors related to the increase in stroke mortality, it is likely multifactorial stemming from both patient factors and disruptions across the continuum of care. From a patient perspective, war-related stress and weakened regulatory oversight has led to a reversal in the decline of smoking^32^, a well-known risk factor for cardiovascular disease. Disruptions across the continuum of care from infrastructure destruction, transportation, and logistics adversely impact patients. This includes both primary care and the treatment of cardiovascular risk factors such as hypertension, as well as the ability to manage acute stroke with optimal medical therapies, such as thrombolytic therapy, which must be given within a short window after the onset of symptoms. Additionally, mobilization of the medical workforce for front-line care may limit the personnel available to care for civilians. Taken together, these findings imply that a broad strategy encompassing targeted public health measures, optimized resource allocation, and reinforced medical logistics is urgently needed to improve civilian outcomes during conflict.

Not surprisingly, age and sex were significant risk factors for in-hospital mortality in our study. Consistent with the literature^33^, age was positively associated with in-hospital mortality. However, while some previous studies^34,35^ have found females to have higher risk of stroke mortality than males, our results showed males had higher mortality risk. One recent study of Ukraine stroke patients from 2025 showed no difference in stroke mortality between males and females^36^, although that study was limited to ischemic stroke only. Our findings regarding stroke type (hemorrhagic vs. ischemic) were also consistent with the literature on stroke mortality^37,38^, showing that patients with hemorrhagic strokes had significantly higher risk of in-hospital mortality, especially for brain stem, multiple localized and intraventricular hemorrhage^39^.

### Strengths and Limitations

The main strength of this study is the use of a large population-based dataset from a national EHR system that included all hospitalizations for stroke events in Ukraine from April 2020 to December 2024. One limitation of the study is the lack of data prior to April 2020. This presents a challenge because the data begin at the same time of the COVID-19 Pandemic, which may have also induced a mortality increase in the pre-war period. Had data been available in the pre-Pandemic period prior to 2020 a better baseline mortality rate could have been established. Unfortunately, the national EHR system was not implemented until early 2020, so the data prior to the Pandemic are not available. Thus, our estimates of excess death may be conservative because they are based on a post-Pandemic baseline. While we attempted to control confounding in our analyses by using multivariable regression and propensity-matching, there is also the potential for residual confounding from unmeasured covariates. Another limitation of the use of EHR data is that data are only recorded if a patient is admitted to the hospital. Thus, this study does not include any minor stroke events that did not involve a hospitalization or any stroke events that involved a prehospital death. Finally, the size and structure of the Ukrainian population has changed over the last few years, which may have also influenced the incidence of stroke.

## CONCLUSION

This is the first study to assess stroke mortality at the population level in Ukraine before and after the start of the full-scale Russo-Ukrainian war and to estimate war-attributable excess mortality. The findings show that in-hospital mortality from stroke events increased after the start of the war, with an estimated 3,944 excess stroke deaths attributable to the war. This study adds to the growing body of evidence that exposure to war leads to indirect population health impacts due to increased risk of chronic disease and mortality, in addition to the direct health impacts from war-related injuries.

### Author Contributions

Dr. Howard had full access to the data in the study and takes responsibility for the integrity of the data and the accuracy of the data analysis.

Study concept and design: OT, JTH

Acquisition, analysis, or interpretation of data: OT, JTH

Drafting of the manuscript: OT, IJS, JTH

Critical revision of the manuscript for important intellectual content: OT, IJS, YH, DL, JTH

Statistical analysis: JTH

Obtained funding:

Administrative, technical, or material support: OT Study supervision: OT

### Conflict of interest Disclosures

All authors have completed and submitted the ICMJE Forms for Disclosure of Potential Conflict of Interest and none were reported.

### Funding/Support

The acquisition of data was sponsored by the World Bank in Ukraine.

### Role of the Funder/Sponsor

The sponsor had no role in the conduct of the research.

### Funding

None

### Previous Presentation or Publication

None

## Data Availability

The data are administered by the Ukraine Ministry of Health and the authors do not have permission to share the data.

## Notes

### Competing Interest Statement

The authors have declared no competing interest.

### Clinical Trial

Not applicable

### Author Declarations

The research was determined to be exempt from review by the University of Texas at San Antonio institutional review board and conducted in accordance with the Declaration of Helsinki and with applicable Ukrainian and U.S. regulations.

